# COVID-19 Time of Intubation Mortality Evaluation (C-TIME): A System for Predicting Mortality of Patients with COVID-19 Pneumonia at the Time They Require Mechanical Ventilation

**DOI:** 10.1101/2022.01.09.22268977

**Authors:** Robert A Raschke, Pooja Rangan, Sumit Agarwal, Suresh Uppalapu, Nehan Sher, Steven C Curry, C. William Heise

**Author notes:** Corresponding author: Robert A Raschke MD, 475 N 5^th^ Street, Phoenix AZ, 85004. Funded in part by a grant from the Flinn Foundation, Phoenix, AZ.

## Abstract

**Background:** An accurate system to predict mortality in patients requiring intubation for COVID-19 could help to inform consent, frame family expectations and assist end-of-life decisions.

**Research objective:** To develop and validate a mortality prediction system called C-TIME (COVID-19 Time of Intubation Mortality Evaluation) using variables available before intubation, determine its discriminant accuracy, and compare it to APACHE IVa and SOFA.

**Methods:** A retrospective cohort was set in 18 medical-surgical ICUs, enrolling consecutive adults, positive by SARS-CoV 2 RNA by reverse transcriptase polymerase chain reaction or positive rapid antigen test, and undergoing endotracheal intubation. All were followed until hospital discharge or death. The combined outcome was hospital mortality or terminal extubation with hospice discharge. Twenty-five clinical and laboratory variables available 48 hours prior to intubation were entered into multiple logistic regression (MLR) and the resulting model was used to predict mortality of validation cohort patients. AUROC was calculated for C-TIME, APACHE IVa and SOFA.

**Results:** The median age of the 2,440 study patients was 66 years; 61.6 percent were men, and 50.5 percent were Hispanic, Native American or African American. Age, gender, COPD, minimum mean arterial pressure, Glasgow Coma scale score, and PaO_2_/FiO_2_ ratio, maximum creatinine and bilirubin, receiving factor Xa inhibitors, days receiving non-invasive respiratory support and days receiving corticosteroids prior to intubation were significantly associated with the outcome variable. The validation cohort comprised 1,179 patients. C-TIME had the highest AUROC of 0.75 (95%CI 0.72-0.79), vs 0.67 (0.64-0.71) and 0.59 (0.55-0.62) for APACHE and SOFA, respectively (Chi^2^ P<0.0001).

**Conclusions:** C-TIME is the only mortality prediction score specifically developed and validated for COVID-19 patients who require mechanical ventilation. It has acceptable discriminant accuracy and goodness-of-fit to assist decision-making just prior to intubation. The C-TIME mortality prediction calculator can be freely accessed on-line at https://phoenixmed.arizona.edu/ctime.

## Introduction

The Coronavirus disease 2019 (COVID-19) pandemic raised concern that an overwhelming surge of critically-ill patients might require exclusion of patients with high predicted mortality from receiving mechanical ventilation (1). The majority of COVID-19 ventilator triage policies surveyed in 2020 incorporated the Sequential Organ Failure Assessment (SOFA) score to predict mortality (2). However, a subsequent study that collected SOFA score data 48 hours prior to intubation yielded a discriminant accuracy for mortality prediction of only 0.59 (95%CI: 0.55-0.63) (3). Although many other scoring systems have been developed to predict mortality in patients with COVID-19 (4-27), none focused on assessing the patient at the time of intubation, when patients, families and providers are forced to make critical decisions regarding life support. Although the capacity to provide mechanical ventilation to COVID-19 patients is improved, the need for ventilator triage is still possible in regional hotspots, and informed consent for endotracheal intubation should include discussion of prognosis. Our aim was to develop a mortality prediction system we called C-TIME (<u>C</u>OVID-19 <u>T</u>ime of Intubation <u>M</u>ortality <u>E</u>valuation) using variables typically available in the 48 hours before intubation, in order to inform consent, frame family expectations and assist end-of-life planning. Our secondary aims were to validate C-TIME, determine its discriminant accuracy and compare it to SOFA and APACHE IVa mortality prediction models.

## Materials and Methods

### Study design

A retrospective cohort study, approved by our IRB, was set in 18 medical surgical ICUs in the Southwest United States between 6/1/2020 and 3/23/2021. June was chosen for cohort inception when preliminary results of the RECOVERY trial (28) were released, and administration of dexamethasone rapidly adopted in our study ICUs. We randomly split our cohort in half to compose model-development and validation cohorts.

### Participants

Consecutive ICU patients were included based on the following eligibility criteria: ≥18 years of age; positive SARS-CoV 2 RNA by reverse transcriptase polymerase chain reaction or positive rapid antigen test; and undergoing endotracheal intubation ≥4 hours after admission. All patients were followed until hospital discharge or death.

### Variables

The main outcome variable was hospital mortality or discharge to hospice after terminal extubation – henceforth this combined outcome is referred to as “mortality”. We chose candidate predictor variables to use in model development based on previous literature (4-27) and hypotheses generated by our clinical research team. We examined our clinical dataset and only selected candidate predictor variables missing in less than 10% of study patients. We made an exception for the partial pressure of arterial oxygen/fraction of inspired oxygen (PaO_2_/FiO_2_) ratio, which we hypothesized would be a particularly important predictor (29); therefore we planned to impute missing PaO_2_/FiO_2_ data (see statistics section below).

The following 25 candidate predictor variables, collected in the time period before intubation, were chosen to include in model development. Patient characteristics included: age, gender, body mass index (BMI), prior history of diabetes mellitus, hypertension, COPD, coronary artery disease, cancer or solid organ transplant. Physical examination findings included maximum temperature, lowest mean arterial pressure and lowest Glasgow Coma scale in the 48 hours prior to intubation. Laboratory variables included the highest concentration of creatinine and bilirubin, and the lowest platelet count and PaO_2_/FiO_2_ ratio in the 48 hours prior to intubation. Management variables comprised hospital days prior to intubation; hospital days receiving non-invasive respiratory support (high-flow nasal canula oxygen; continuous positive airway pressure or bilevel positive airway pressure) prior to intubation; hospital days receiving corticosteroids (dexamethasone, methylprednisolone or prednisone) prior to intubation; and administration of any of the following drugs: corticosteroids, therapeutic dose heparin/enoxaparin, oral Xa inhibitors, subcutaneous or intravenous insulin, or norepinephrine infusion. We also included intubation during surge conditions, defined as the time period(s) during which ≥ 400 ventilators (>5.5 ventilators per 100,000 population) were in use by COVID-19 patients in the state of Arizona where most of our study hospitals were located. By this criteria, surge conditions occurred in our ICUs in the summer (6/23/2020 - 8/7/2020) and winter (12/3/2020 - 2/14/2021) (30). Variables needed to calculate the SOFA score were also extracted in the 48 hours prior to intubation and used to derive the associated predicted mortality for each patient (31,32). Variables used to calculate APACHE IVa predicted mortality were obtained in the first 24 hours after ICU admission, as the APACHE system is designed to operate.

### Data sources

We extracted and de-identified clinical data from a Cerner Millenium® electronic medical record (EMR). Our hospital system uses the proprietary APACHE IVa® severity scoring system (Cerner Corp, Kansas City MO) to calculate predicted hospital mortality for each ICU patient that meets criteria for APACHE calculations (33).

### Study size

We calculated that a sample size of 2500 patients would allow analysis of 25 candidate predictor variables in our logistic regression - one variable for every 50 patients in the model-development and validation cohorts of 1,250 patients, each.

### Statistical Analysis

All study patients underwent randomization into two equal-sized model-development and validation cohorts. Missing FiO_2_ values were imputed as the mean FiO_2_ for all study patients for whom FiO_2_ was known. Missing PaO_2_ values were imputed as the mean PaO_2_ of all study patients receiving the same FiO_2_. The 25 candidate predictor variables were entered into backwards, step-wise, multiple logistic regression (MLR) using the model-development cohort, with mortality as the dependent outcome variable. We retained all variables that remained in the model at P≤0.05.

The MLR logistic equation from the model development cohort was then applied to calculate predicted mortality for each patient in the validation cohort and to calculate Nagelkerke’s pseudo R^2^ and Hosmer-Lemeshow goodness-of-fit tests. We compared goodness-of-fit and sensitivity for C-TIME versus SOFA and APACHE IVa in patients with very high predicted mortality, for whom prognostic information is most likely to affect end-of-life decisions. The three models were each used to identify clinically important patient subgroups with ≥75%, ≥90% and ≥95% predicted mortality, and the *observed* mortality proportion in each of the resulting subgroups was enumerated. The number of observed deaths in each subgroup over the total number of observed deaths in the validation cohort equaled the sensitivity of each model at each of the three predicted mortality cutoffs. We also looked at observed mortality proportion for patients C-TIME predicted to have <50% mortality. The area under the receiver operator characteristic curve (AUROC), a measurement of discriminant accuracy, for C-TIME, SOFA and APACHE IVa were calculated and compared using the Chi-squared statistic. The Wilson method was used to calculate 95% confidence intervals for single proportions. We used STATA^®^ Version 17 (Statacorp, College Station, TX) for all statistical analyses.

## Sensitivity analysis

We also calculated AUROCs for C-TIME and SOFA using only validation cohort patients for whom FiO_2_ and PaO_2_ were known (i.e. *excluding* patients with imputed values). These were compared to the AUROCs of our primary analysis by using z-tests on equality of proportions to test whether data imputation affected AUROC.

### Results

Between 6/1/2020 and 3/23/2021, 18,431 patients with COVID-19 were admitted to study hospitals. Of these, 4,695 were admitted to the ICU and 2,440 were intubated ≥4 hours after admission. Characteristics of these 2,440 study patients are presented in the table 1. The median age was 66 years, 61.6 percent were men, and 50.5 percent were Hispanic, Native American or African American. Eighty-six percent of patients received corticosteroids. Eleven variables were significant in the final MLR model (see table 2). The validation cohort comprised 1,219 patients of whom 1,179 had complete data for analysis by MLR. Observed mortality in the validation cohort was 65.1%.

**Table 1:**
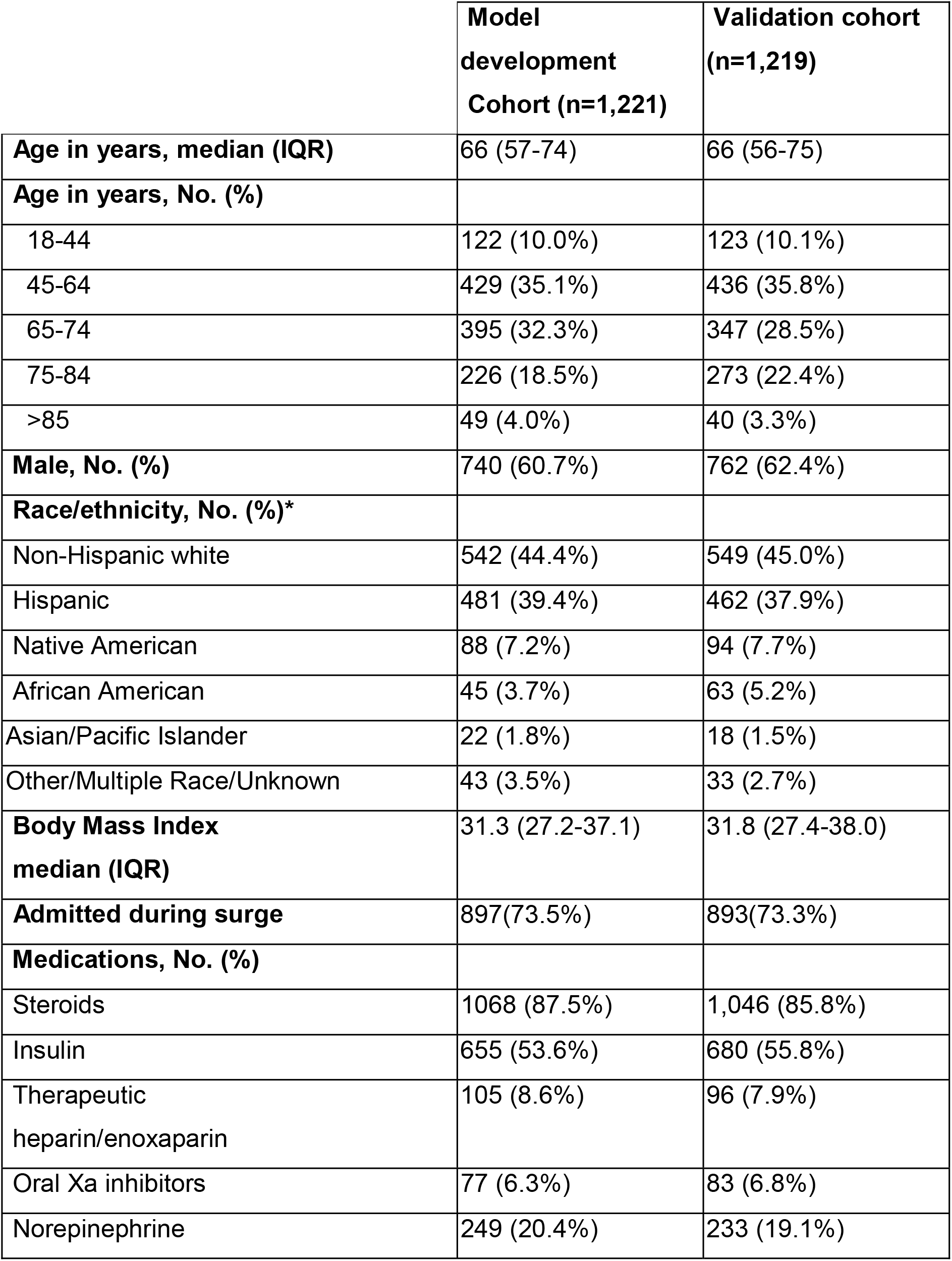
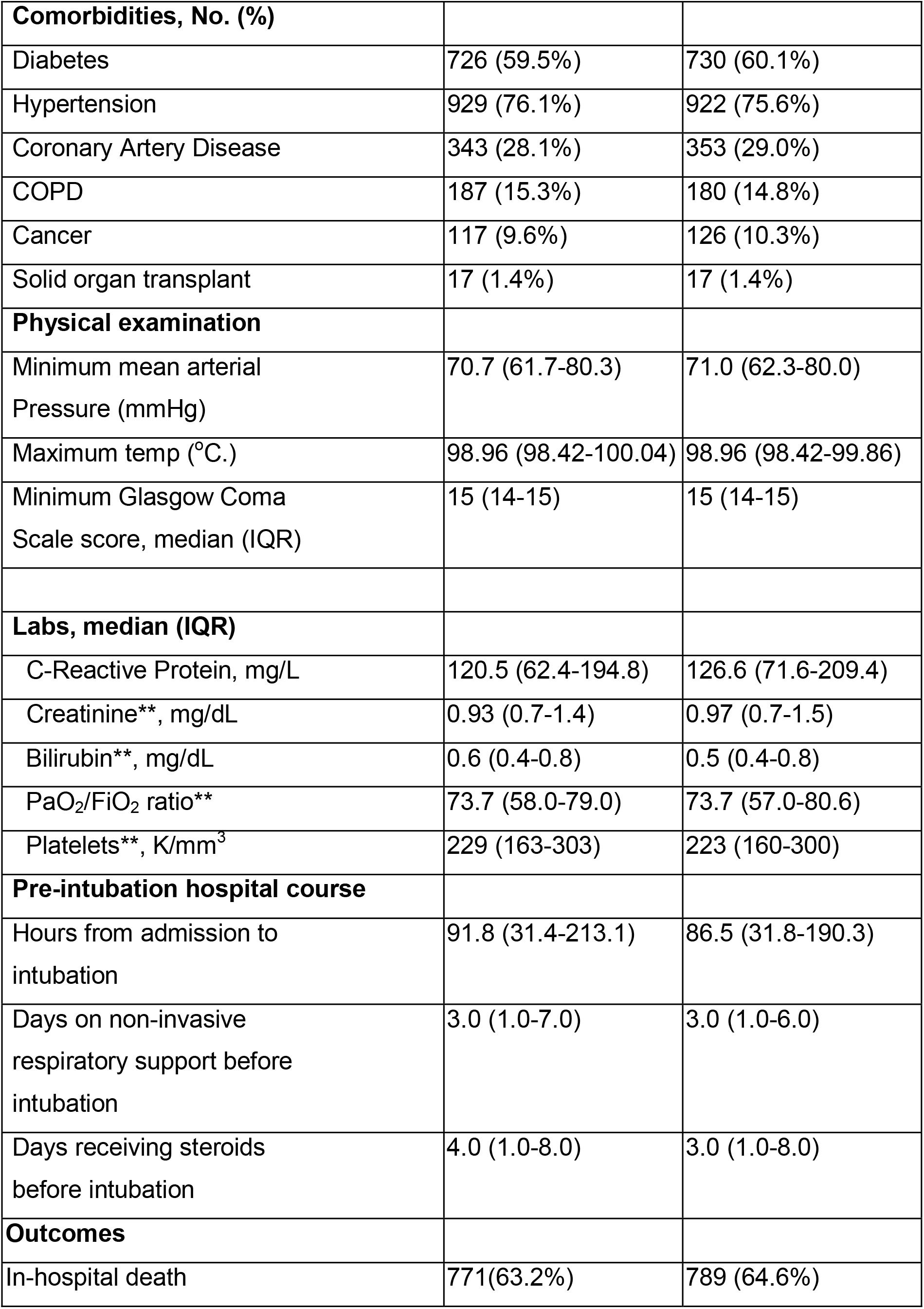
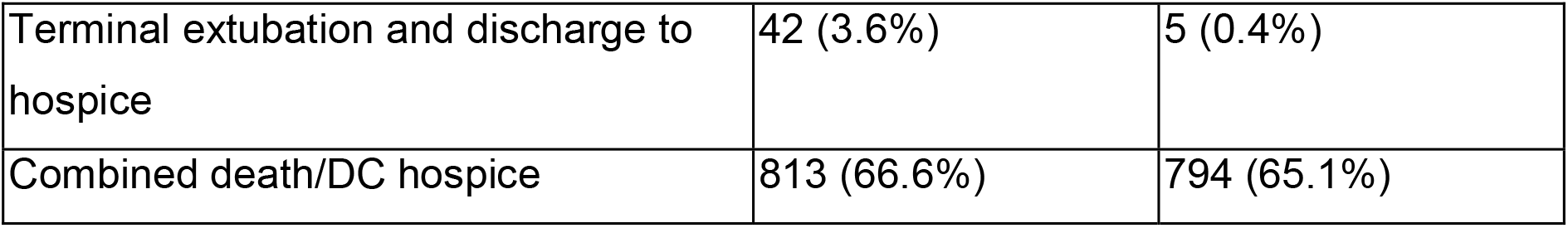
Clinical characteristics of 2440 study patients.

|  | **Model**<br>**development**<br>**Cohort (n=1,221)** | **Validation cohort**<br>**(n=1,219)** |
| --- | --- | --- |
| **Age in years, median (IQR)** | 66 (57-74) | 66 (56-75) |
| **Age in years, No. (%)** |  |  |
| 18-44 | 122 (10.0%) | 123 (10.1%) |
| 45-64 | 429 (35.1%) | 436 (35.8%) |
| 65-74 | 395 (32.3%) | 347 (28.5%) |
| 75-84 | 226 (18.5%) | 273 (22.4%) |
| >85 | 49 (4.0%) | 40 (3.3%) |
| **Male, No. (%)** | 740 (60.7%) | 762 (62.4%) |
| **Race/ethnicity, No. (%)\*** |  |  |
| Non-Hispanic white | 542 (44.4%) | 549 (45.0%) |
| Hispanic | 481 (39.4%) | 462 (37.9%) |
| Native American | 88 (7.2%) | 94 (7.7%) |
| African American | 45 (3.7%) | 63 (5.2%) |
| Asian/Pacific Islander | 22 (1.8%) | 18 (1.5%) |
| Other/Multiple Race/Unknown | 43 (3.5%) | 33 (2.7%) |
| **Body Mass Index**<br>**median (IQR)** | 31.3 (27.2-37.1) | 31.8 (27.4-38.0) |
| **Admitted during surge** | 897(73.5%) | 893(73.3%) |
| **Medications, No. (%)** |  |  |
| Steroids | 1068 (87.5%) | 1,046 (85.8%) |
| Insulin | 655 (53.6%) | 680 (55.8%) |
| Therapeutic<br>heparin/enoxaparin | 105 (8.6%) | 96 (7.9%) |
| Oral Xa inhibitors | 77 (6.3%) | 83 (6.8%) |
| Norepinephrine | 249 (20.4%) | 233 (19.1%) |
| **Comorbidities, No. (%)** |  |  |
| Diabetes | 726 (59.5%) | 730 (60.1%) |
| Hypertension | 929 (76.1%) | 922 (75.6%) |
| Coronary Artery Disease | 343 (28.1%) | 353 (29.0%) |
| COPD | 187 (15.3%) | 180 (14.8%) |
| Cancer | 117 (9.6%) | 126 (10.3%) |
| Solid organ transplant | 17 (1.4%) | 17 (1.4%) |
| **Physical examination** |  |  |
| Minimum mean arterial Pressure (mmHg) | 70.7 (61.7-80.3) | 71.0 (62.3-80.0) |
| Maximum temp (°C.) | 98.96 (98.42-100.04) | 98.96 (98.42-99.86) |
| Minimum Glasgow Coma Scale score, median (IQR) | 15 (14-15) | 15 (14-15) |
| **Labs, median (IQR)** |  |  |
| C-Reactive Protein, mg/L | 120.5 (62.4-194.8) | 126.6 (71.6-209.4) |
| Creatinine**, mg/dL | 0.93 (0.7-1.4) | 0.97 (0.7-1.5) |
| Bilirubin**, mg/dL | 0.6 (0.4-0.8) | 0.5 (0.4-0.8) |
| PaO<sub>2</sub>/FiO<sub>2</sub> ratio** | 73.7 (58.0-79.0) | 73.7 (57.0-80.6) |
| Platelets**, K/mm<sup>3</sup> | 229 (163-303) | 223 (160-300) |
| **Pre-intubation hospital course** |  |  |
| Hours from admission to intubation | 91.8 (31.4-213.1) | 86.5 (31.8-190.3) |
| Days on non-invasive respiratory support before intubation | 3.0 (1.0-7.0) | 3.0 (1.0-6.0) |
| Days receiving steroids before intubation | 4.0 (1.0-8.0) | 3.0 (1.0-8.0) |
| **Outcomes** |  |  |
| In-hospital death | 771(63.2%) | 789 (64.6%) |
| Terminal extubation and discharge to hospice | 42 (3.6%) | 5 (0.4%) |
| Combined death/DC hospice | 813 (66.6%) | 794 (65.1%) |

**Table 2:** Multiple Logistic Regression model with significant predictor variables for the outcome mortality in the model-development cohort.

| **Significant predictor variables in the C-TIME MLR model:** | **Odds ratio\* (95% CI)** | **P value** |
| --- | --- | --- |
| **Age (years)** | 1.71 (1.47-1.98) | <0.001 |
| **Male Gender** | 1.41 (1.06-1.89) | 0.019 |
| **COPD** | 1.63 (1.07-2.49) | 0.024 |
| **Minimum mean arterial pressure (mmHg)** | 0.81 (0.70-0.93) | 0.004 |
| **Minimum Glasgow Coma Scale score** | 0.82 (0.70-0.95) | 0.008 |
| **PaO<sub>2</sub>/FiO<sub>2</sub> (mmHg)** | 0.73 (0.62-0.86) | <0.001 |
| **Maximum creatinine (mg/dl)** | 1.17 (1.00-1.36) | 0.050 |
| **Maximum bilirubin (mg/dl)** | 1.55 (1.13-2.13) | 0.006 |
| **Days receiving non-invasive respiratory support** | 1.52 (1.08-2.13) | 0.017 |
| **Days receiving corticosteroids** | 1.43 (1.05-1.94) | 0.024 |
| **Received oral Xa inhibitors** | 2.37 (1.16-4.85) | 0.018 |
| *Odds ratios are associated with a one standard deviation (SD) increment for continuous variables. Values used for SD: age 13.7 years; MAP 13.7 mmHg; PaO<sub>2</sub>/FiO<sub>2</sub> 78.3 mmHg; creatinine 1.9 mg/dl; bilirubin 2.0 mg/dl, days receiving corticosteroids 5 days; Minimum Glasgow Coma Scale score 3; Days receiving non-invasive respiratory support before intubation 5 days. |  |  |

C-TIME AUROC was 0.75 (95%CI 0.72-0.79), Nagelkerke’s pseudo R^2^ = 0.25, and the Hosmer-Lemeshow Chi^2^ showed acceptable goodness-of-fit with P = 0.29 in the validation cohort. C-TIME classified 486 (26.3%), 141 (11.6%) and 43 (3.5%) validation cohort patients as having ≥75%, ≥90% and ≥95% predicted mortality respectively. *Observed* mortality in these three subgroups was 82% (95%CI: 78-85%), 91% (95%CI: 85-95%) and 95% (95%CI: 85-99%), respectively. In contrast, APACHE IVa classified 46 (3.8%), 15 (1.2%) and 5 (0.4%) patients as having ≥75%, ≥90% and ≥95% predicted mortality, respectively. Observed mortality in the three APACHE IVa subgroups was 89% (95%CI: 77-95%), 80% (9F%CI: 55-93%) and 100% (95%CI 57-100%). The highest SOFA score achieved predicted mortality of 89%; therefore SOFA did not predict any patient to have >90% mortality (see table 3). C-TIME also classified 246/1179 (21%) of patients as having ≤50% mortality – these patients had a mean predicted mortality of 32.1% and an observed mortality of 35.7%.

**Table 3:** Observed mortality proportions for three subgroups of patients with high predicted mortality (≥75, ≥90 and ≥95% predicted mortality) identified by three mortality prediction systems.

| | Patients with $\geq 75\%$<br>Predicted mortality | Patients with $\geq 90\%$<br>Predicted mortality | Patients with $\geq 95\%$<br>Predicted mortality |
| --- | --- | --- | --- |
| C-TIME | 399/486<br>82%<br>(95%CI: 78-85%) | 128/141<br>91%<br>(95%CI: 85-95%) | 41/43<br>95%<br>(95%CI: 85-99%) |
| APACHE IVa | 41/46<br>89%<br>(95%CI: 77-95%) | 12/15<br>80%<br>(95%CI: 55-93%) | 5/5<br>100%<br>(95%CI: 57-100%) |
| SOFA | 92/120<br>77%<br>(95%CI: 68-83%) | NA | NA |
| NA: not applicable – the highest SOFA score was associated with predicted mortality of 89% |  |  |  |

Nine-hundred-sixty-two patients in our validation cohort had met criteria for APACHE IVa calculations and were included in our comparison of AUROC between C-TIME, APACHE IVa and SOFA. C-TIME had the highest AUROC of 0.75 (95%CI 0.72-0.79), vs 0.67 (0.64-0.71) and 0.59 (0.55-0.62) for APACHE and SOFA, respectively (Chi^2^ P<0.0001). See figure.

**Figure legend.**
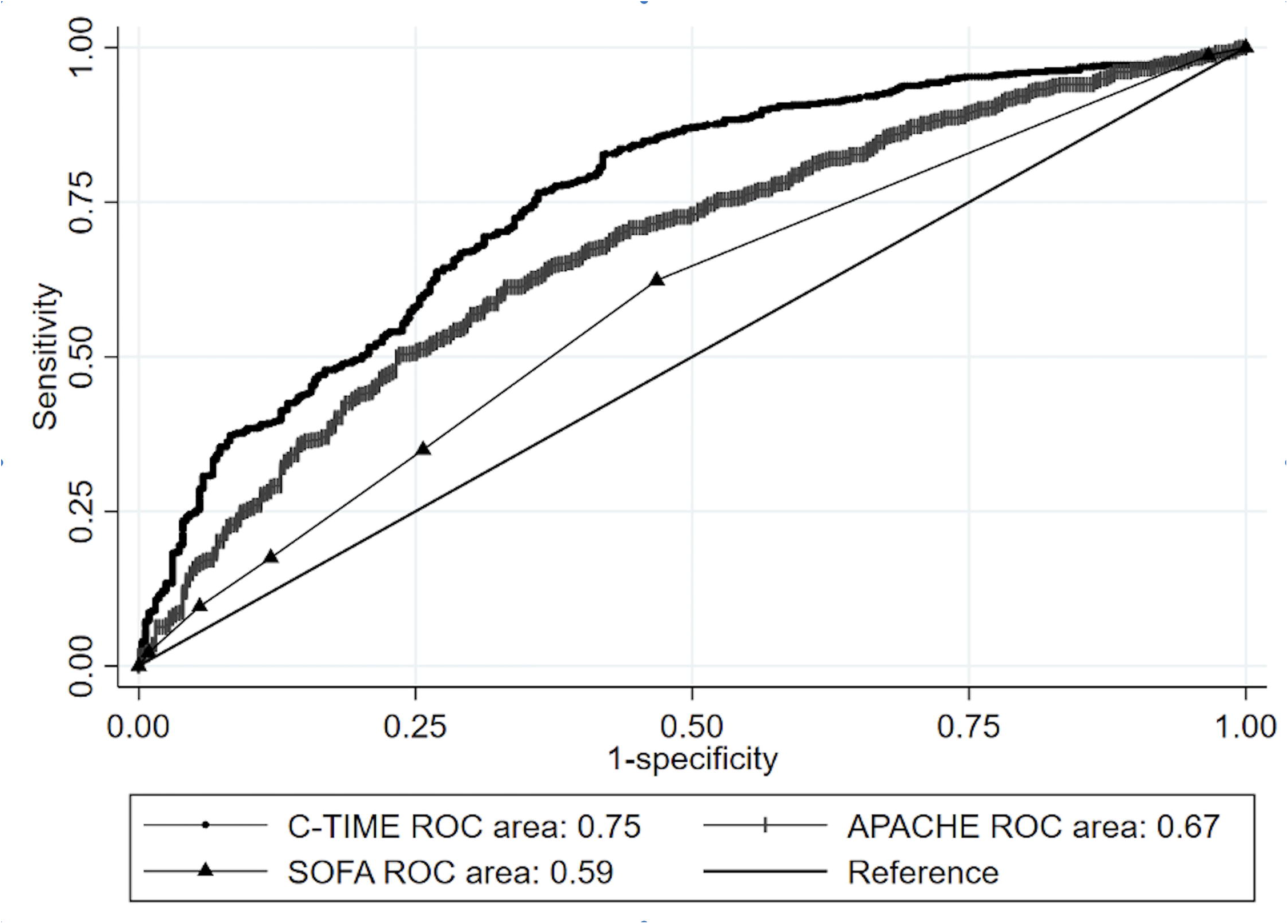
Comparative AUROC of C-TIME, APACHE IVa, and SOFA mortality prediction systems.

### Sensitivity analysis in relationship to imputed data

FiO_2_ was imputed to be 95.6% in 202 patients (8.3%). This is unlikely to have introduced bias since 1950/2240 (87%) of study patients for whom FiO_2_ was recorded had an FiO_2_ of 100%. PaO_2_ was imputed in 647 cases (26.5%). Sensitivity analysis showed that C-TIME and SOFA AUROCs in the subset of validation patients *without* imputed PaO_2_ were 0.75 (CI 0.71-0.79) and 0.58 (CI 0.54-0.62) respectively – essentially identical to AUROCs calculated for the full validation cohort.

## Discussion

The C-TIME mortality prediction model, based on eleven easily obtained clinical and laboratory variables, has better discriminant accuracy than APACHE IVa with 145 variables (33). Furthermore, the C-TIME model has acceptable “goodness of fit” and sensitivity in patients with high predicted mortality, in whom C-TIME may be helpful in making end-of-life decisions. Our study hospitals range from tertiary academic centers to community and critical access facilities serving a variety of persons from urban and rural communities with a wide diversity of racial/ethnic backgrounds and socioeconomic status, enhancing the external generalizability of our findings.

Well over one hundred prognostic systems, including general ICU systems (such as SOFA and APACHE), and novel systems specifically developed for COVID-19 patients have already been published to predict clinical outcomes in patients with COVID-19 (4). These vary by target patient population, predictor variables and outcomes of interest. To provide context for C-TIME, we reviewed comparable scoring systems that were developed and validated specifically for hospitalized COVID-19 patients and which incorporated commonly available clinical and laboratory predictor variables, and which reported AUROCs for in-hospital mortality (5-27).

Several features distinguish C-TIME from other validated COVID-19 mortality prediction systems we reviewed. 1) C-TIME is the only system that specifically evaluates patients with COVID-19 pneumonia just before they require mechanical ventilation. The discriminant accuracy of other prognostic models at this point in a patient’s clinical course are unknown, due to spectrum effect: “When designing a predictive scoring system, the study cohort should be as similar as possible to the population in which the test is intended to be used” (34). 2) The C-TIME study cohort had by far the highest reported mortality (65%) of any of the previous studies, as would be expected for *intubated* COVID-19 patients (35). The mortality of the study cohort has a strong influence on the operating characteristics of associated mortality prediction systems (34) – another reason why previously reported mortality prediction scores are likely not reliable if used at the time of intubation. 3) Other mortality prediction systems utilized study cohorts that included patients admitted prior to 6/2020, when preliminary results of RECOVERY were released. The inclusion of significant numbers of patients who did not receive corticosteroids could limit their generalizability in relationship to current practice patterns. Eighty-six percent of our study patients received corticosteroids before intubation. 4) C-TIME is the only model that incorporates treatment variables. Days receiving corticosteroids and days receiving non-invasive respiratory support prior to intubation were associated with mortality in our model-development and validation cohorts, and were also significantly associated with surge conditions (p=0.0003 and 0.005, respectively). Surviving patients received a median of two days steroids and two days non-invasive respiratory support; non-survivors received a median of nine days steroids and eight days non-invasive respiratory support. A recent study showed that mortality increased significantly during the winter COVID-19 surge (36) however a meta-analysis concluded that delaying intubation does *not* influence mortality (37). It is possible that the associations observed in our study might be due to prolonged efforts at non-invasive respiratory support and corticosteroid treatment of patients during surge conditions, selecting treatment non-responders for intubation. Inclusion of these treatment variables associated with surge conditions suggests that C-TIME might retain generalizability regardless of whether or not surge conditions prevail in the clinical setting in which it is used.

One particular C-TIME variable deserves brief comment. We tested pre-intubation antithrombotic therapy with heparin, enoxaparin and factor Xa inhibitors in model development based on the hypothesis that these drugs might prevent fatal thromboembolic complications related to COVID-19. However, therapeutic heparin/enoxaparin fell out of the model, and receiving factor Xa inhibitors was surprisingly associated with *increased* mortality. Analysis of a sample of these patients showed that pre-existing atrial fibrillation was the indication for factor Xa inhibitors in 80% of patients. It is possible that receiving a factor Xa inhibitor was a confounding variable representing pre-existing atrial fibrillation in our model.

Several other mortality prediction systems with acceptable operating characteristics are available for prognostication at the *time of admission* to the hospital, rather than at the time of intubation. The 4C score is supported by the largest study cohort and has an AUROC (0.77) similar to C-TIME (5). We noted the highest AUROCs were reported for systems that prognosticated using variables from the time of admission in Hubei province early in the pandemic (8,14,18,23,25). We feel these results are likely irreproducible outside the special circumstances under which they were reported. This contention is supported by a study using data from the Veterans Affairs Data Warehouse (11) that externally-validated several of these prediction scores (23,25) and found much lower AUROCs than those originally reported: 0.68 vs. 0.91, 0.72 vs. 0.94, respectively. This phenomenon was also demonstrated for the SOFA score, which achieved AUROCs of 0.89 (0.83-0.96) (39) and 0.99 (0.98-1.00) (40) in Hubei province early in the pandemic, versus 0.58 and 0.61 (0.53-0.70) in larger, more recent studies from the US and UK (3,5).

Examination of Table 3 reveals that C-TIME has good fit when it predicts very high mortality, and that it is more sensitive than APACHE IVa or SOFA at identifying non-survivors. There were 399 fatal outcomes among patients predicted by C-TIME to have ≥75% predicted mortality, yielding a sensitivity of 399/794 (50.3%). The comparative sensitivity for APACHE IVa was 41/794 (5.2%) and for SOFA was 92/794 (11.6%). Sensitivity quantifies the potential clinical impact that C-TIME could have should resuscitative measures be limited due to high predicted mortality.

### Limitations of the study

Missing data was a major complication of our retrospective cohort design that limited us from including less-frequently-ordered predictor variables such as C-reactive protein, and led us to impute missing PaO_2_ and FiO_2_ data. Our sensitivity analysis showed that the later did not affect our AUROC estimates. Our EMR data source limited our ability to include variables not recorded as discrete data, such as COVID-19 vaccination status and pre-existing atrial fibrillation.

The discriminant accuracy achieved by C-TIME was modest, although similar to several other COVID-19 mortality prediction systems with AUROCs ranging 0.72-0.79 (5,10,17,19,21,38). We believe that it is inherently difficult to predict COVID-19 mortality at the time of intubation because such patients are relatively clinical homogeneous; most have life-threatening, single organ, respiratory failure (see table 1) (3). Low variation in predictor variables reduces discriminant accuracy. This could explain why APACHE IVa, which achieved AUROC of 0.88 in a large general ICU population (33), only yielded an AUROC of 0.66 in our study cohort.

C-TIME (and all other COVID-19 prognostic systems) are likely to lose discriminant accuracy over time, as factors influencing survival evolve. These factors might include advances in therapy and emergence of new viral strains. The aforementioned decline in discriminant accuracy reported in Hubei vs the US and UK demonstrate that discriminant accuracy demonstrated in one historical juncture is likely not generalizable in future clinical settings. Thus, any prognostic scoring system for COVID-19 will likely require repeated validation over time.

## Conclusions

C-TIME is the only currently available mortality predictive score specifically developed and validated for COVID-19 patients who require intubation. It has acceptable discriminant accuracy and goodness-of-fit to assist informed consent for intubation and other end-of-life issues that occur specifically at this critical juncture in the patient’s care. The C-TIME predicted mortality calculator can be accessed free on-line at: https://phoenixmed.arizona.edu/ctime

## Data Availability

All data produced in the present study are available upon reasonable request to the authors

## Abbreviations

APACHE: acute physiology and chronic health evaluation
AUROC: area under the receiver-operating curve
BMI: body mass index
COPD: chronic obstructive pulmonary disease
COVID-19: coronavirus disease 2019
C-TIME: COVID-19 time of intubation mortality evaluation
FiO2: fraction of inspired oxygen
MLR: multiple logistic regression
PaO2: partial pressure of arterial oxygen
SARS CoV-2: severe acute respiratory syndrome coronavirus 2
SOFA: sequential organ failure assessment

## Guarantor statement

Dr Raschke had full access to all of the data in the study and takes personal responsibility for the integrity of the data and the accuracy of the data analysis.

## Author Contributions

*Concept and design:* Raschke, Rangan, Agarwal, Uppalapu, Sher, Curry and Heise.*Acquisition, analysis, or interpretation of data:* Agarwal, Sher *Drafting of the manuscript:* Raschke.

*Critical revision of the manuscript for important intellectual content:* Rangan, Agarwal, Uppalapu, Sher, Curry and Heise.

*Statistical analysis:* Rangan.

*Obtained funding:* Curry.

*Supervision:* Curry.

The authors have no conflicts of interest to declare

This research was performed with partial support from a Flinn Foundation grant, but the Flinn Foundation had no influence over the conduct of the study, interpretation of the data, preparation of the manuscript or the decision to submit for publication.

